# Glycated Protein Precipitation Index: Bridging the Gap Between Short-Term and Long-Term Glycemic Markers

**DOI:** 10.64898/2025.12.12.25342172

**Authors:** Luís Jesuino de Oliveira Andrade, Gabriela Correia Matos de Oliveira, Luís Matos de Oliveira

**Affiliations:** Department of Health of the State University of Santa Cruz – Ilhéus – Bahia – Brazil.; José Silveira Foundation – Salvador – Bahia – Brazil

**Keywords:** Glycated proteins, Diabetes mellitus, Glycemic monitoring, Point-of-care diagnostics

## Abstract

**Background:** Current glycemic monitoring exhibits a critical temporal gap between fructosamine (14-21 days) and HbA1c (90-120 days), limiting timely therapeutic adjustments. The Glycated Protein Precipitation Index (GPPI) represents a novel biomarker potentially bridging this interval.

**Objective:** To validate GPPI’s correlation with established glycemic markers and determine its clinical utility for intermediate-term glycemic assessment. Methods: This prospective validation study will enroll 200 diabetic patients and 100 normoglycemic controls. GPPI leverages differential precipitation of glycated versus native serum proteins using trichloroacetic acid at optimized concentrations, followed by colorimetric quantification with bromophenol blue at 595nm. Primary endpoint: correlation coefficient between GPPI and HbA1c (target r>0.85). Secondary endpoints include analytical precision (CV<5%), interference assessment, reference interval establishment, and cost-effectiveness analysis. Simultaneous HbA1c, fasting glucose, and continuous glucose monitoring (subset n=50) will provide comparative data. Deming regression, Bland-Altman analysis, and ROC curves will assess method agreement and diagnostic performance.

**Expected Outcomes:** GPPI should reflect 6-8 week glycemic control with strong HbA1c correlation, offering cost reduction from $15-30 to $0.24 per test. Simplified visual interpretation enables field deployment in resource-limited settings, potentially democratizing diabetes monitoring for underserved populations globally.

## INTRODUCTION

Diabetes mellitus (DM) affects approximately 589 million adults worldwide, with projections indicating this will reach 853 million by 2050.^1^ This chronic metabolic disorder, characterized by sustained hyperglycemia, precipitates devastating microvascular complications including retinopathy, nephropathy, and neuropathy, alongside macrovascular sequelae such as coronary heart disease and stroke.^2^ The pathophysiological mechanisms underlying these complications involve chronic hyperglycemia, oxidative stress, advanced glycation end products, and endothelial dysfunction, underscoring the critical importance of maintaining optimal glycemic control throughout the disease trajectory.^3^

Accurate assessment of glycemic status remains fundamental to diabetes management, with hemoglobin A1c (HbA1c) serving as the cornerstone biomarker, reflecting average blood glucose over approximately 8 to 12 weeks.^4^ Despite its widespread utility and validation in major clinical trials, HbA1c exhibits important limitations in conditions affecting erythrocyte turnover, including anemias, hemoglobinopathies, chronic kidney disease, and pregnancy.^5^ Alternative markers such as fructosamine, reflecting 2 to 3 weeks of glycemic exposure, provide complementary assessment when HbA1c interpretation proves problematic.^6^

A critical gap exists between short-term markers like fructosamine and long-term indicators such as HbA1c. This intermediate timeframe of 4 to 8 weeks represents a clinically significant period during which therapeutic decisions are frequently made, including medication adjustments and evaluation of newly initiated treatments. Current monitoring strategies lack a validated biomarker optimally suited for this window, forcing clinicians to either wait for delayed HbA1c changes or rely on sporadic self-monitoring data. This gap becomes particularly problematic in pregnancy, evaluation of therapeutic interventions requiring prompt feedback, and populations where HbA1c reliability is compromised.

Drawing upon the chemistry of protein glycation, this study aims to introduce a novel laboratory methodology designed to assess the average glycemic levels over an 8-week period, thereby bridging the gap between existing glycemic markers.

## PROJECT METHODOLOGY

### 1. Project Overview

#### 1.1 Rationale

Current glycemic monitoring in resource-limited settings faces significant barriers: HbA1c testing requires sophisticated equipment and trained specialists, while fructosamine assays demand refrigerated reagents and consistent electrical supply. This methodology proposes validation of the Glycated Protein Precipitation Index (GPPI), that leverages differential solubility properties of glycated versus non-glycated serum proteins when exposed to trichloroacetic acid (TCA).

#### 1.2 Scientific Foundation

Glycated serum proteins exhibit altered physicochemical properties, including modified electrostatic charge distribution and tertiary structure. When treated with TCA at 7.5% concentration, glycated proteins precipitate preferentially compared to native proteins, enabling simple quantification through colorimetric detection following differential precipitation.

#### 1.3 Temporal Window

GPPI reflects average glycemic control over 6-8 weeks (42-56 days), positioning it between fructosamine (14-21 days) and HbA1c (90-120 days), potentially offering an optimal monitoring interval for treatment adjustment.

### 2. Research Objectives

#### 2.1 Primary Objective

To validate the correlation between GPPI and established glycemic markers (HbA1c, mean plasma glucose) in a heterogeneous diabetic population across diverse clinical settings.

#### 2.2 Secondary Objectives

- Establish reference ranges for normoglycemic, prediabetic, and diabetic populations
- Determine analytical precision, accuracy, and reproducibility
- Assess interference from common confounding factors (hemolysis, lipemia, hypoalbuminemia)
- Develop and validate simplified visual colorimetric interpretation for field deployment
- Calculate cost-effectiveness compared to standard monitoring approaches
- Design implementation protocols for resource-limited healthcare facilities

### 3. Experimental Design

#### 3.1 Study Population

***Phase 1 – Method Development (n=50)***

- Pilot cohort for optimization of TCA concentrations and incubation parameters
- Equal distribution: 25 diabetic patients, 25 normoglycemic controls
- Inclusion: Age 18-75 years, confirmed diagnosis (diabetics), fasting glucose <100 mg/dL (controls)

***Phase 2 – Validation Study (n=300)***

- 200 patients with established diabetes mellitus (Type 1 and Type 2)
- 100 normoglycemic controls
- Stratified by glycemic control level based on recent HbA1c:
  ➢ Excellent control: HbA1c <7.0% (n=50)
  ➢ Moderate control: HbA1c 7.0-9.0% (n=100)
  ➢ Poor control: HbA1c >9.0% (n=50)
  ➢ Controls: HbA1c <5.7% (n=100)

**Inclusion Criteria:**

- Adults aged 18-75 years
- Confirmed diabetes diagnosis >6 months (diabetic cohort)
- No acute illness requiring hospitalization within past 30 days
- Willing to provide informed consent

**Exclusion Criteria:**

- Severe anemia (hemoglobin <8 g/dL)
- Advanced chronic kidney disease (eGFR <30 mL/min/1.73m²)
- Active malignancy or immunosuppressive therapy
- Pregnancy or lactation
- Multiple myeloma or paraproteinemia
- Severe hepatic dysfunction (total bilirubin >3× ULN)
- Recent blood transfusion (<120 days)

### 4. Laboratory Methodology

#### 4.1 Reagent Preparation

***Stock Solutions:***

- **Trichloroacetic Acid (TCA) Solutions**
  ➢ TCA 15% (w/v): 150g TCA dissolved in deionized water to 1000mL
  ➢ TCA 7.5% (w/v): 75g TCA dissolved in deionized water to 1000mL
  ➢ Storage: Amber glass bottles, room temperature, 12-month stability
- **Phosphate Buffer 0.1M, pH 7.4**
  ➢ Na□HPO□·7H□O: 26.8g
  ➢ NaH□PO□·H□O: 13.8g
  ➢ Deionized water to 1000mL
  ➢ Adjust pH to 7.4 ± 0.1 using 1N NaOH or 1N HCl
  ➢ Storage: Room temperature, 6-month stability
- **Bromophenol Blue 0.04% (w/v)**
  ➢ 40mg bromophenol blue powder
  ➢ 100mL deionized water
  ➢ Storage: Amber bottle, room temperature, 6-month stability
- Sodium Hydroxide 0.1N
  ➢ 4g NaOH pellets
  ➢ Deionized water to 1000mL
  ➢ Storage: Plastic bottle, room temperature, 12-month stability

#### 4.2 Sample Collection and Processing

***Specimen Type:*** Venous blood, serum

***Collection Volume:*** 3-5mL whole blood

***Collection Tube:*** Red-top tube (no anticoagulant) or serum separator tube (SST)

**Processing Protocol**:

1. Allow blood to clot at room temperature: 30-45 minutes
2. Centrifuge at 1500-2000 × g for 10 minutes
3. Carefully separate serum without disturbing cellular interface
4. Transfer to labeled polypropylene tube
5. If not analyzed immediately: Store at 2-8°C (up to 72 hours) or –20°C (extended storage)

**Pre-analytical Quality Checks**:

- Visual inspection for hemolysis, icterus, lipemia
- Document any specimen abnormalities
- Reject grossly hemolyzed specimens (hemolysis index >200 mg/dL hemoglobin)

#### 4.3 GPPI Analytical Procedure

**Equipment Required**:

- Calibrated spectrophotometer with 595nm filter (±2nm bandwidth)
- Temperature-controlled centrifuge (2000 × g capacity)
- Calibrated adjustable micropipettes (100-5000 μL range)
- Borosilicate glass test tubes (13×100mm) or polypropylene equivalents
- 37°C water bath or incubator
- Vortex mixer
- Timer

**Standardized Protocol – Duplicate Determination**:

**TUBE A (Total Protein Precipitation)**:

1. Pipette 0.5mL serum into labeled tube
2. Add 4.5mL phosphate buffer (0.1M, pH 7.4)
3. Mix gently by inversion (3-5 times)
4. Add 5.0mL TCA 15%
5. Mix immediately but gently (avoid foaming)
6. Incubate at room temperature (20-25°C) for 10 minutes ± 30 seconds
7. Centrifuge at 2000 × g for 5 minutes
8. Carefully decant supernatant (discard)
9. Invert tube on absorbent paper for 30 seconds to drain residual liquid
10. Add 5.0mL NaOH 0.1N to precipitate
11. Vortex vigorously for 30 seconds to completely resuspend precipitate
12. Add 0.2mL bromophenol blue 0.04%
13. Mix well by inversion
14. Incubate at room temperature for 5 minutes
15. Read absorbance at 595nm against deionized water blank
16. Record as A□ (total protein absorbance)

TUBE B (Glycated Protein Precipitation):

1. Pipette 0.5mL serum into labeled tube
2. Add 4.5mL phosphate buffer (0.1M, pH 7.4)
3. Mix gently by inversion (3-5 times)
4. Add 2.5mL TCA 7.5% ← Critical difference: lower TCA concentration
5. Mix immediately but gently (avoid foaming)
6. Incubate at room temperature (20-25°C) for 10 minutes ± 30 seconds
7. Centrifuge at 2000 × g for 5 minutes
8. Carefully decant supernatant (discard)
9. Invert tube on absorbent paper for 30 seconds to drain residual liquid
10. Add 5.0mL NaOH 0.1N to precipitate
11. Vortex vigorously for 30 seconds to completely resuspend precipitate
12. Add 0.2mL bromophenol blue 0.04%
13. Mix well by inversion
14. Incubate at room temperature for 5 minutes
15. Read absorbance at 595nm against deionized water blank
16. Record as A□ (glycated protein absorbance)

**Calculation:**

***GPPI (%) = (A□ / A□) × 100***

Where:

A□ = Absorbance from Tube B (glycated protein fraction)

A□ = Absorbance from Tube A (total protein fraction)

**Estimated Mean Plasma Glucose (mg/dL):**

***MPG = (GPPI × 5.2) + 16***

This formula provides correlation with 6-8 week average glucose levels.

#### 4.4 Quality Control Procedures

**Daily QC Protocol**:

***Level 1 – Normal Control (Target GPPI: 12-18%)***

- Pooled serum from normoglycemic volunteers
- Aliquoted and frozen at –80°C
- Thaw one aliquot daily, use within 8 hours

***Level 2 – Diabetic Control (Target GPPI: 35-45%)***

- Pooled serum from poorly controlled diabetic patients
- Aliquoted and frozen at –80°C
- Thaw one aliquot daily, use within 8 hours

***Level 3 – Extreme Control (Target GPPI: >50%)***

- Normal serum spiked with glucose (500 mg/dL)
- Incubated at 37°C for 24 hours to induce glycation
- Aliquoted and frozen at –80°C

**Acceptance Criteria**:

- QC values must fall within ±2 SD of established mean
- If QC fails: Troubleshoot (reagent integrity, equipment calibration, technique)
- Document all QC results in laboratory logbook

**Weekly Calibration**:

- Spectrophotometer wavelength accuracy verification (Holmium oxide filter)
- Linearity check using bromophenol blue serial dilutions
- Centrifuge speed verification (tachometer)

### 5. Comparative Reference Methods

#### 5.1 HbA1c Determination

- Method: High-Performance Liquid Chromatography (HPLC) or immunoturbidimetric assay
- Specimen: EDTA whole blood
- Timing: Collected simultaneously with serum sample for GPPI
- Laboratory: NGSP-certified laboratory
- Target: ≤72-hour turnaround

#### 5.2 Fasting Plasma Glucose

- Method: Hexokinase enzymatic assay
- Specimen: Sodium fluoride/potassium oxalate whole blood
- Timing: Fasting state (≥8 hours)
- Analysis: Within 30 minutes of collection

#### 5.3 Continuous Glucose Monitoring (Subset Analysis)

- Device: Professional CGM system (FreeStyle Libre Pro)
- Duration: 14 days prior to blood collection
- Subset: 50 diabetic patients
- Metric: Time in range, mean glucose, glycemic variability

#### 5.4 Additional Biochemical Parameters

- Serum albumin (bromocresol green method)
- Serum creatinine (enzymatic method)
- Complete blood count with hemoglobin
- Lipid profile (triglycerides, total cholesterol, HDL and LDL cholesterol)

### 6. Analytical Validation Parameters

#### 6.1 Precision Studies

***Within-Run Precision (Intra-assay CV)***:

- 20 replicates of 3 QC levels analyzed in single run
- Calculate mean, SD, CV%
- Acceptance: CV <5% for all levels

***Between-Run Precision (Inter-assay CV)***:

- 3 QC levels analyzed in duplicate over 20 consecutive days
- Calculate mean, SD, CV%
- Acceptance: CV <8% for all levels

***Inter-Operator Precision***:

- 3 trained technicians each perform 10 replicates of 3 QC levels
- ANOVA analysis for operator effect
- Acceptance: No significant operator bias (p>0.05)

#### 6.2 Accuracy Studies

**Method Comparison**:

- Deming regression: GPPI vs HbA1c (n=200)
- Calculate slope, intercept, correlation coefficient (r)
- Bland-Altman plot for bias assessment
- Target: r >0.85, systematic bias <10%

**Recovery Studies**:

- Spike known glycated protein standards into serum
- Expected spike concentrations: 10%, 20%, 30% increase in GPPI
- Calculate percent recovery
- Acceptance: 90-110% recovery

**Conversion Formula Validation**:

- GPPI = (HbA1c × 6.2) – 6
- Compare predicted GPPI vs measured GPPI
- Calculate prediction error
- Assess clinical agreement using Clarke Error Grid

#### 6.3 Analytical Sensitivity

**Limit of Detection (LoD)**:

- Analyze 20 replicates of normal serum (lowest expected GPPI)
- Calculate mean + 3SD
- Establish minimum detectable GPPI value

**Limit of Quantitation (LoQ)**:

- Lowest GPPI concentration with CV <10%
- Serial dilution of high GPPI sample

**Analytical Measuring Range**:

- Linearity assessment from 10% to 60% GPPI
- Serial dilution of high GPPI samples
- Polynomial regression analysis

#### 6.4 Specificity and Interference

**Studies Hemolysis Interference**:

- Prepare samples with increasing hemoglobin concentrations (50, 100, 200, 500 mg/dL)
- Compare GPPI to non-hemolyzed control
- Document interference threshold

**Lipemia Interference**:

- Add Intralipid to create triglyceride levels: 200, 500, 1000, 2000 mg/dL
- Assess GPPI deviation from control
- Establish acceptable lipemia limit

**Icterus Interference**:

- Spike samples with bilirubin: 5, 10, 20, 40 mg/dL
- Evaluate impact on GPPI measurement
- Define bilirubin interference threshold

**Hypoalbuminemia**:

- Stratify patients by albumin level (<2.5, 2.5-3.5, >3.5 g/dL)
- Assess GPPI values in each stratum
- Develop correction factor if needed

**Paraproteinemia**:

- Test samples from patients with monoclonal gammopathies
- Compare to matched diabetic controls
- Establish exclusion criteria

#### 6.5 Stability Studies

**Room Temperature Stability:**

- Store serum at 20-25°C
- Test GPPI at 0, 4, 8, 24, 48, 72 hours
- Define maximum storage time (≤10% deviation)

**Refrigerated Stability**:

- Store serum at 2-8°C
- Test GPPI at 0, 3, 7, 14, 21 days
- Establish refrigerated shelf life

**Frozen Stability**:

- Store serum at –20°C and –80°C
- Test GPPI at 1, 3, 6, 12 months
- Evaluate freeze-thaw cycles (1-5 cycles)

**Reagent Stability**:

- Monitor all reagents monthly over 18-month period
- Perform GPPI on QC samples
- Document reagent expiration recommendations

### 7. Clinical Validation

#### 7.1 Reference Interval Establishment

**Normoglycemic Population (n=100):**

- CLSI EP28-A3c guidelines
- Age-stratified: 18-40, 41-60, >60 years
- Gender-balanced
- Exclusion of outliers (Dixon’s Q-test)
- Nonparametric 95% reference interval (2.5th-97.5th percentile)

**Prediabetic Range**:

- Individuals with impaired fasting glucose (100-125 mg/dL)
- HbA1c 5.7-6.4%
- n=50 for preliminary range

#### 7.2 Diagnostic Performance

**ROC Curve Analysis**:

- GPPI discrimination between normoglycemia and diabetes
- Calculate AUC, optimal cutoff, sensitivity, specificity
- Compare to HbA1c threshold (6.5%)

**Clinical Concordance**:

- Classify patients using GPPI-derived ranges vs HbA1c-based classification
- Calculate kappa statistic for agreement
- Misclassification rate analysis

**Longitudinal Tracking (Subset n=50):**

- Serial GPPI measurements every 6 weeks × 6 months
- Parallel HbA1c every 12 weeks
- Assess tracking of glycemic changes
- Correlation with therapeutic interventions

#### 7.3 Clinical Decision-Making Impact

**Proposed GPPI Reference Ranges**:

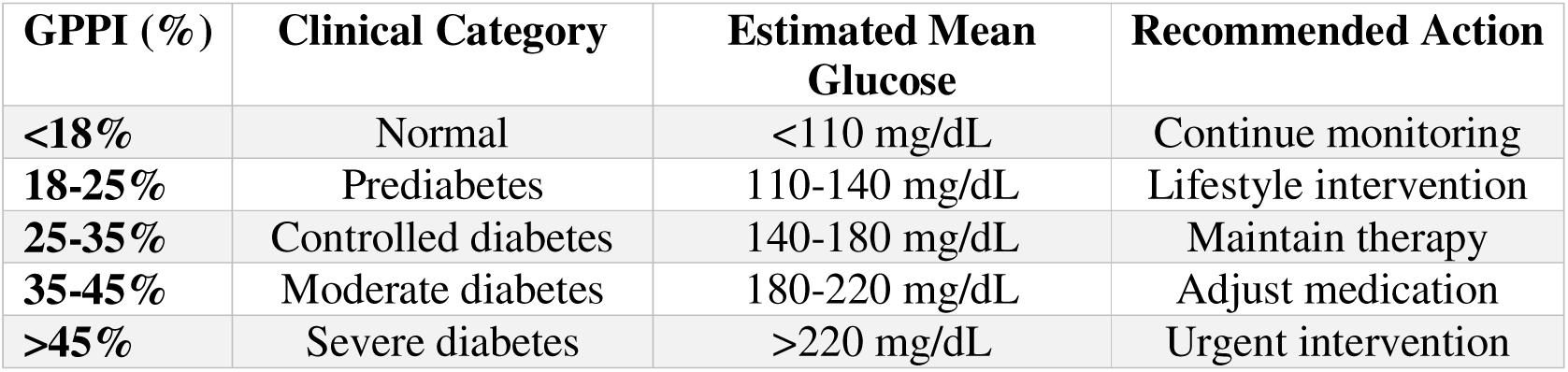

| GPPI (%) | Clinical Category | Estimated Mean Glucose | Recommended Action |
| --- | --- | --- | --- |
| <18% | Normal | <110 mg/dL | Continue monitoring |
| 18-25% | Prediabetes | 110-140 mg/dL | Lifestyle intervention |
| 25-35% | Controlled diabetes | 140-180 mg/dL | Maintain therapy |
| 35-45% | Moderate diabetes | 180-220 mg/dL | Adjust medication |
| >45% | Severe diabetes | >220 mg/dL | Urgent intervention |

**Validation:**

- Prospective assessment of therapeutic decisions based on GPPI
- Compare outcomes to HbA1c-guided management
- Patient satisfaction surveys

### 8. Simplified Visual Method Development

#### 8.1 Colorimetric Scale Creation

**Objective:** Field-deployable interpretation without spectrophotometer

**Methodology**:

1. Prepare GPPI standards spanning 10-60% range (5% increments)
2. Process using standard protocol through bromophenol blue addition
3. Professional color photography under standardized lighting (D65 illuminant)
4. Print on laminated waterproof cards
5. Include Tube A reference (100% intensity = dark blue)

**Visual Scale Interpretation**:

- Very light blue (10-15%): Normal
- Light blue (15-25%): Prediabetes
- Medium blue (25-35%): Controlled diabetes
- Dark blue (35-45%): Moderate diabetes
- Very dark blue (>45%): Severe diabetes

#### 8.2 Visual Method Validation

**Observer Study (n=30 operators):**

- 20 blinded GPPI samples per operator
- Compare visual estimation vs spectrophotometric measurement
- Calculate correlation, mean absolute error
- Target: ±8% agreement for 90% of readings

**Field Testing**:

- 5 rural health clinics without reliable electricity
- 50 patient samples per clinic
- Visual interpretation by trained health workers
- Correlation with reference laboratory GPPI

### 9. Implementation Feasibility Assessment

#### 9.1 Cost Analysis

**Per-Test Direct Costs:**

- TCA 15%: $0.05
- TCA 7.5%: $0.03
- Phosphate buffer: $0.02
- Bromophenol blue: $0.03
- NaOH: $0.01
- Consumables (tubes, pipettes): $0.10
- Total per test: $0.24

**Capital Equipment (Basic Laboratory)**:

- Spectrophotometer: $300-800
- Centrifuge: $200-500
- Pipettes: $150-300
- Water bath: $100-200
- Total equipment: $750-1,800

#### 9.2 Training Requirements

**Tier 1 – Basic Technician Training:**

- Duration: 4 hours (theory + hands-on)
- Competency: Perform GPPI protocol, basic QC
- Certification: Pass practical examination (5 consecutive successful QC runs)

**Tier 2 – Laboratory Supervisor Training:**

- Duration: 8 hours
- Competency: Troubleshooting, QC documentation, result interpretation
- Certification: Written exam + practical assessment

**Training Materials:**

- Illustrated protocol manual (Portuguese/Spanish/English)
- Video demonstrations (15 minutes)
- Quick reference laminated cards
- QC logbook templates

#### 9.3 Scalability Assessment

**Phase 1 – Proof of Concept (Month 1-3):**

- Single reference laboratory
- Complete analytical validation (n=300 samples)
- Protocol optimization
- Budget: $5,000

**Phase 2 – Regional Pilot (Month 4-6):**

- 10 primary health centers
- Train 20 technicians
- Process 1,000 patient samples
- Logistics testing (reagent distribution, result reporting)
- Budget: $15,000

**Phase 3 – Scale-Up (Month 7-12):**

- 100 health facilities
- 5,000 patients tested monthly
- Establish central quality oversight
- Data management system
- Budget: $75,000

### 10. Data Management and Statistical Analysis

#### 10.1 Data Collection

**Electronic Data Capture:**

**REDCap or equivalent secure database**

- **Variables collected:**
  ➢ Demographics (age, gender, ethnicity, BMI)
  ➢ Diabetes type, duration, current medications
  ➢ GPPI results (A□, A□, calculated GPPI, repeat measurements)
  ➢ HbA1c, fasting glucose, CGM metrics
  ➢ Albumin, creatinine, hemoglobin
  ➢ Specimen quality indicators (hemolysis, lipemia, icterus)
  ➢ Operator ID, date, laboratory site

#### 10.2 Statistical Methods

**Correlation Analysis:**

- Pearson correlation: GPPI vs HbA1c, fasting glucose, CGM mean
- Spearman correlation for non-normal distributions
- 95% confidence intervals for correlation coefficients

**Method Comparison:**

- Deming regression (accounts for measurement error in both methods)
- Bland-Altman analysis (bias and limits of agreement)
- Passing-Bablok regression (non-parametric alternative)

**Diagnostic Accuracy:**

- ROC curve analysis with AUC calculation
- Optimal cutoff determination (Youden index)
- Sensitivity, specificity, PPV, NPV at various thresholds
- Likelihood ratios

**Subgroup Analysis:**

- Stratification by diabetes type, duration, glycemic control
- ANOVA for between-group comparisons
- Multivariate regression for confounding adjustment

**Reproducibility:**

- Intraclass correlation coefficient (ICC)
- Coefficient of variation (CV)
- Bland-Altman repeatability coefficient

#### 10.3 Sample Size and Power

**Primary Endpoint (Correlation):**

- Null hypothesis: r = 0.70 (moderate correlation)
- Alternative hypothesis: r = 0.90 (strong correlation)
- Power: 90%, α = 0.05 (two-tailed)
- Required n: 156 subjects
- Accounting for 20% attrition: n = 195 → Target n = 200

**Secondary Endpoint (Diagnostic Accuracy):**

- Expected AUC = 0.90
- Null AUC = 0.75
- Power: 80%, α = 0.05
- Diabetic: Control ratio 2:1
- Required n: 180 (120 diabetic, 60 control)
- Covered by primary endpoint sample size

### 11. Ethical Considerations

#### 11.1 Regulatory Approval

- Institutional Review Board (IRB) submission and approval
- Clinical trial registration (ClinicalTrials.gov or equivalent)
- Informed consent process in local language
- Confidentiality and data protection (HIPAA or local equivalent)

#### 11.2 Participant Protection

- Voluntary participation, right to withdraw
- No deviation from standard care based solely on GPPI results during validation
- All participants receive standard HbA1c testing
- Reporting of critical values to treating physician
- Adverse event monitoring and reporting

#### 11.3 Benefit-Risk Assessment

- Minimal risk: Standard phlebotomy
- Direct benefit: Comprehensive glycemic assessment
- Societal benefit: Potential for improved diabetes monitoring in underserved areas

### 12. Timeline and Milestones

**Month 1-2: Protocol Development and IRB Approval**

- Finalize study protocol
- Prepare informed consent documents
- Submit IRB application
- Procure equipment and reagents

**Month 3: Method Optimization (Phase 1)**

- Pilot testing (n=50)
- Optimize TCA concentrations
- Refine incubation times
- Initial precision studies

**Month 4-6: Analytical Validation**

- Precision studies (within-run, between-run, inter-operator)
- Accuracy and recovery experiments
- Interference testing
- Stability assessments
- Establish QC procedures

**Month 7-9: Clinical Validation (Phase 2)**

- Enroll 200 diabetic patients + 100 controls
- Simultaneous GPPI, HbA1c, fasting glucose measurement
- CGM substudy (n=50)
- Complete data collection

**Month 10-11: Data Analysis and Manuscript Preparation**

- Statistical analysis
- Visual method validation
- Cost-effectiveness modeling
- Draft scientific manuscript

**Month 12: Dissemination and Implementation Planning**

- Present results at scientific conference
- Submit manuscript for peer review
- Develop implementation toolkit
- Train pilot site personnel

### 13. Expected Outcomes and Deliverables

#### 13.1 Primary Deliverables

***1.*** ***Validated GPPI Methodology***
  - Standardized protocol document
  - Reference range establishment
  - QC procedures and acceptance criteria
***2.*** ***Correlation Equation***
  - GPPI-to-HbA1c conversion formula with 95% CI
  - GPPI-to-mean glucose estimation with validation
***3.*** ***Simplified Visual Interpretation Tool***
  - Colorimetric reference card
  - Field validation data
**4.** **Implementation Package**
  - Training manual (illustrated, multilingual)
  - Equipment procurement guide
  - Cost analysis spreadsheet
  - QC logbooks and forms

### 14. Quality Assurance

#### 14.1 Internal QC

- Daily three-level QC (normal, diabetic, extreme)
- Levey-Jennings charts for trend monitoring
- Westgard rules for QC violation detection
- Monthly reagent lot verification

#### 14.2 External Quality Assessment

- Participation in interlaboratory comparison program
- Quarterly proficiency testing
- Peer laboratory exchange program
- Blind duplicate testing (10% of samples)

#### 14.3 Documentation

- Master protocol with version control
- Standard Operating Procedures (SOPs) for each analytical step
- Equipment maintenance logs
- Training records and competency assessments
- Deviation and corrective action reports

### 15. Risk Management and Mitigation

| Risk | Probability | Impact | Mitigation Strategy |
| --- | --- | --- | --- |
| Poor GPPI-HbA1c correlation | Low | High | Thorough method optimization in Phase 1; adjust TCA concentrations if needed |
| Recruitment challenges | Medium | Medium | Engage multiple clinical sites; extend enrollment period if necessary |
| Interference from common conditions | Medium | Medium | Comprehensive interference testing; develop correction algorithms |
| Reagent supply chain issues | Low | Low | Source from multiple vendors; validate alternative reagent brands |
| Equipment failure | Low | Medium | Backup equipment; establish repair/replacement protocols |
| Data quality issues | Low | High | REDCap validation rules; regular data audits; centralized monitoring |
| Regulatory delays | Medium | Low | Early IRB engagement; pre-submission meetings; maintain clear communication |

### 16. Sustainability and Long-Term Vision

#### 16.1 Commercialization Pathway

- Develop standardized GPPI kit for distribution
- Pursue regulatory clearance (ANVISA)
- Partner with diagnostic manufacturers for scale-up
- Establish supply chain in target countries

#### 16.2 Capacity Building

- Train-the-trainer programs for regional expansion
- Integration into laboratory medicine curricula
- Certification programs for GPPI technicians
- Continuous medical education (CME) for clinicians

#### 16.3 Research Extensions

- Pediatric population validation
- Gestational diabetes application
- Non-diabetic hyperglycemia conditions
- Combination with point-of-care glucose meters
- Digital connectivity (smartphone colorimetry)

## CONCLUSION

This methodology outlines a rigorous, phased approach to validate the Glycated Protein Precipitation Index as a practical alternative to HbA1c for glycemic monitoring in resource-limited settings. The protocol balances scientific rigor with pragmatic feasibility, addressing analytical performance, clinical utility, and implementation barriers. Successful validation could democratize diabetes monitoring for millions of underserved patients globally, representing a paradigm shift in accessible diagnostics.

## Data Availability

All data produced in the present work are contained in the manuscript

